# Vaccine waning and immune escape drive the second surge of Omicron spread in Hong Kong: A modeling study

**DOI:** 10.1101/2023.02.14.23285831

**Authors:** Yuling Zou, Wing-Cheong Lo, Wai-Kit Ming, Hsiang-Yu Yuan

## Abstract

The Omicron subvariant BA.2 caused the start of the fifth epidemic wave in Hong Kong in early 2022, leading to a significant outbreak and triggering more people get immunized in a population with relatively low vaccine coverage. About half a year later, a second outbreak, largely dominated by BA.4 and BA.5 subvariants, began to spread, which peaked within few months. How the waning of the vaccine protection and the immune escape properties together with other factors, including social distancing and outdoor temperature, drove the second surge of infections is unknown. The challenge is that basic epidemic modeling is not able to capture longitudinal change in vaccine waning and dose interval. We developed mathematical equations to formulate continuous change of vaccine waning after observing empirical serological data and incorporate them into a multi-strain discrete-time SEIR (Susceptible-Exposed-Infectious-Removed) model. Using the reported cases during the first outbreak as the training set together with daily vaccination rates, population mobility and temperature, the model successfully predicted the second surge and the replacement by BA.4/5, leading to a cumulative number of cases about 543,600 (7.27% of total population). If vaccine protection maintained without decreasing, the number of predicted cumulative cases reduced 35%. If perfect vaccine coverage reached (100%) by 1^st^ June, the number of cases was only partially reduced 18.65%. Moderate level of social distancing (10% reduction in population mobility) reduced only 13.78% cases, which was not able to prevent the second surge. The results suggest future local outbreaks will remain as long as new immune escape happens with waning immunity, even if the moderate level of social distancing is present. Therefore, a more accurate model forecasting considering waning immunity remains needed in order to allow a better preparation of the next outbreaks.

## Introduction

COVID-19 has been spreading around the world since 2020. Even though vaccines provide protection against COVID-19 transmission and reduce the number of hospital admissions, the end of a public health emergency is still not yet [1]. Uncertainties remain, for example, the protection from the vaccine declines over time because of waning immunity [2-4] and the virus is still able to evolve to escape from immunity. Therefore, repeated outbreaks may likely to happen. How these factors along with social distancing and outdoor temperature drive the new surges of infections and whether high vaccine coverage is able to prevent future outbreaks are still not clear.

Hong Kong faced a significant health burden from the fifth wave since Chinese New Year in 2022 for about three months and experienced a second surge about half a year later. Omicron (BA.2) first entered the community in December 2021, triggering the outbreak. Up to April 2022, about infected cases were reported (in a total population of 7.48 million) and the highest daily number of new cases exceeds 60,000 [5]. After the peak of the fifth wave, the epidemic was effectively suppressed between April and June (e.g. the number of daily cases was consistently below 1,000). However, a second surge began around July and peaked in September. This wave might be controlled by the combined effects of enlarging social distance and improving vaccine coverage.

During this period, BA.4 and BA.5 (denoted as BA.4/5) subvariants were identified in Hong Kong since June 2022. By the end of October 2022, BA.2 declined while BA.4/5 mutant viruses predominated, occupying about 90% [6]. A previous study has found that BA.4/5 have a greater potential to escape vaccine protection and are substantially (4.2-fold) more resistant to neutralization by sera than BA.2 and thus more likely to lead to vaccine breakthrough infections [7].

During the early fifth wave (between late December 2021 and early March 2022), most people in Hong Kong have completed at least one dose of vaccine and began to receive second or boosting doses. As of January 15th, 2022, the vaccine coverage of residents in Hong Kong is as follows: the first dose was 65.88%, the second dose was 62.93% and the rate of boosting dose was only 5.33%. Two brands of vaccines (BNT162b2 and SINOVAC) were available to the public since February 2021. Both vaccines differed in their level of protection and the trend of waning. A study found that the booster of BNT162b2 provided a higher level of protection but the protection from both vaccines waned significantly within 6 months [8]. 77% protection was found if people received 3 doses of BNT162b2 at 180th days since the second injection but 3 doses of SINOVAC only remained at 8% protection. The protections after two doses of the vaccine (regardless of which brand) are generally very low. Waning protection from high to lower levels allows populations to be re-exposed to high-risk infections along with new subvariants, but only very few studies have quantified the impact of vaccine waning on the new infection surges.

Although many recent studies focus on assessing different vaccine allocation or dosing interval strategies [9,10] using the Susceptible-Exposed-Infectious-Removed (SEIR) model considering vaccination or the waning of immunity, these studies assumed the protection of vaccine fixed at different fixed levels before or after the immunity waned. A long-term continuous change in immunity provides a more accurate description of real immunological data but this has not been incorporated into the SEIR model in recent studies.

The study aimed at forecasting a subsequent outbreak that was caused by new Omicron subvariants when the vaccine-induced immunity continued to decline. We built an SEIR model embedding the daily change in immunity after vaccination and trained the model during the first outbreak. The parameterized model successfully predicted the rise of the second surge after BA.4/5 were imported. The model helps to make better preparation of the next outbreak.

## Methods

Methods were divided into two parts. First, we used sera data to simulate the mathematical function of vaccine protection. Second, we estimated a generalized SEIR combining with time-discrete vaccinated population.

### Vaccine waning immunity

We developed the mathematical function of the effect of boosting and waning vaccine-induced immunity with the following assumptions: First, individuals received the second dose of vaccine 30 days after the first dose on average and the third dose 200 days after the second dose on average. Second, vaccine protection changed non-linearly over time. Thus, we used a generalized exponential function to fit the decay of protection from sera data [11]. Third, the protection increased and peaked on 6th days after vaccination, and then, was followed by the waning [12]. Besides, no matter which brand of vaccine, one dose of vaccine did not provide protection [13]. The mathematical function would be used in later modeling simulation.

### Mathematical modeling

#### Formulation of transmission rate

The time-varying parameter *β* are determined by several factors, including weather, population mobility, and the percentage of vaccinated population. Therefore we have:

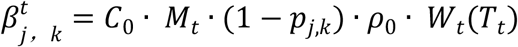

where *j* is the number of vaccines already administered and *k* is the number of days after the last dose. *C*_0_ is the average contact number [14]; *M*_*t*_ is the daily average mobility rate from google website **(Supplementary Figure S1A)** [15] ; *p*_*j,k*_ is the Vaccine protection when an individual has received *j* dose after *k* day; *ρ*_0_ is the reference probability of successful transmission under each contact; and *W*_*t*_ is the nonlinear function of daily temperature *T*_*t*_. *W*_*t*_ was influenced by weather, which is a function defined by daily average temperature **(Supplementary Figure S1B)**. Within a certain range, the increase or decrease of temperature would both enhance the transmission ability of disease [16].

### Discrete-time simulation with vaccination

A discrete-time model with vaccination was developed based on SEIR framework [17-22]. The model assumed that the whole population was initially susceptible (*S* group) and there would be partial population from *S* group received the first dose of vaccine every day. People who had received the first dose of the vaccine and never been infected would receive the second dose (with the probability of *pro*_2_) 30 days (on average) after the injection of first dose. Similarly, people who had received the second dose of the vaccine and never been infected would receive the third dose (with the probability of *pro*_*3*_) 200 days after the first dose (on average). The individuals who had received vaccine and never been infected were defined as vaccinated group (*V* group). Some people in *S* group would be exposed to the virus and become exposed (*E*) group before they were vaccinated. *S* and *V* population become *E* with *β*. β_0_ and *β*_*v*_ are respectively the ability of disease transmission across the susceptible and vaccinated population. After a period of incubation time (1/*δ*), they were detected to be Infectious (*I*) group, and then recovered (*R*). *γ* _0_ is the recovered rate of individuals without vaccine [23] and *β*_*v*_ is the recovered rate with vaccine [24] ; Vaccine protection waned over time. Depending on the number of doses and the days since injection, the protections (*p*_*j,k*_) against the virus are different between different *V*_*j,k*_. The simplified model schematic was plotted (**Fig.1)**. The detailed model schematic and steps can be found in Supplementary Methods and **Fig. S2**).

### Multi-strain modelling

We estimated the impact of BA.4 and BA.5 on protection using viral titre data. Our research only used BNT162b2’s data to replace two brands when focusing on BA.4/5. In order to formulate 3-dose vaccine protection for BA.4/5, we adopted a negative exponential dose–response model [21]. The probability of infection for a susceptible individual is defined as:

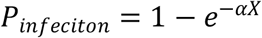

where *X* is numbers of SARS-CoV-2 RNA copies of one susceptible individual and *α* is infectivity. Then the protection is defined as:

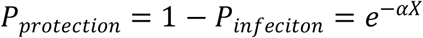

Last, the following is the mathematical relationship between vaccine protection for BA.4/5 and for BA.2 (**Fig. 3D**):

**Fig. 1.**
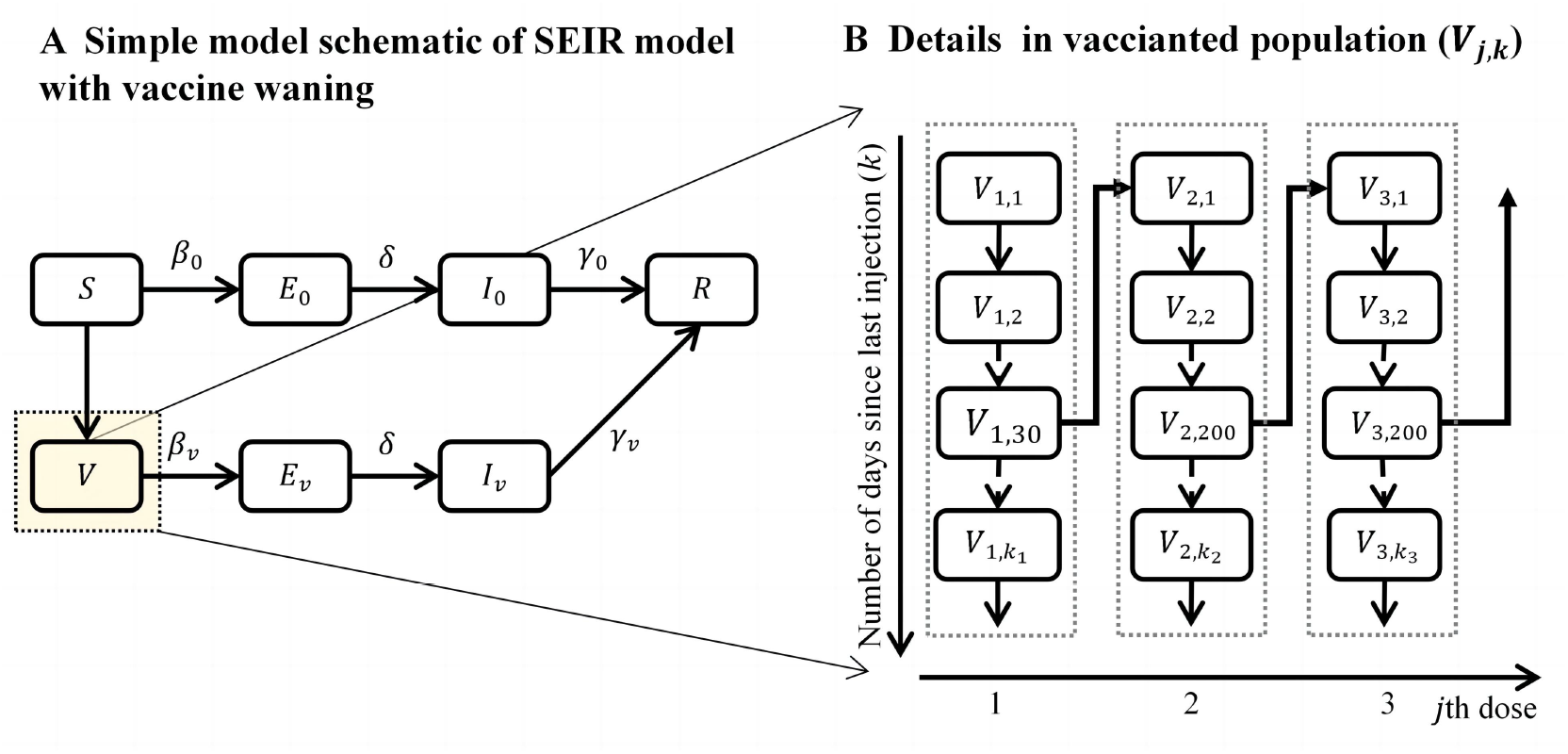
Daily time-step discrete SEIR model with vaccination. Individuals were classified into the following group: *S* (Susceptible, without any vaccination and never been infected), *V* (Individuals with vaccination), *E* (exposed but not yet infected individuals), *I* (infected individuals, including pre-clinical, clinical, and sub-clinical infections) and *R* (recovered, including hospital confirmed, recovered and removed individuals).

**Fig. 2.**
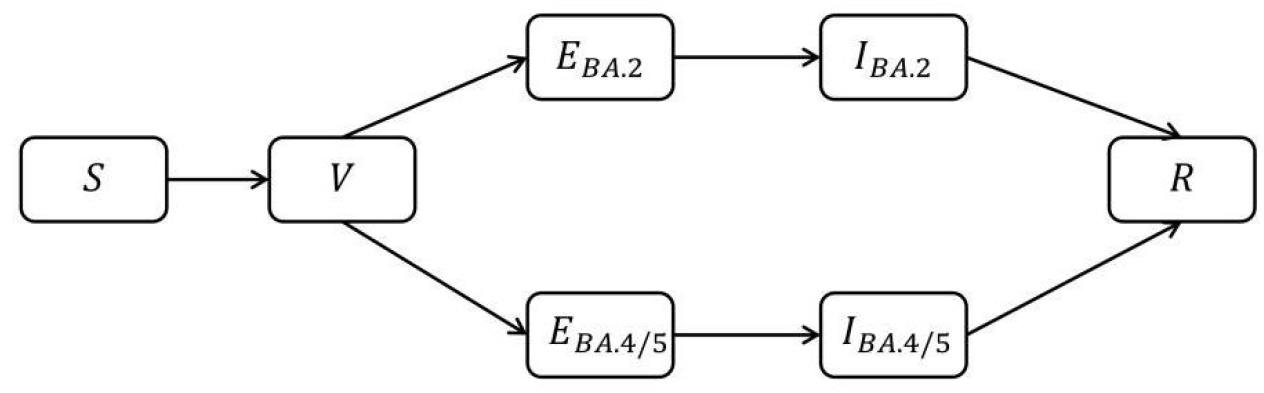
Daily time-step discrete SEIR model with vaccination for cross-immunity.

**Fig. 3.**
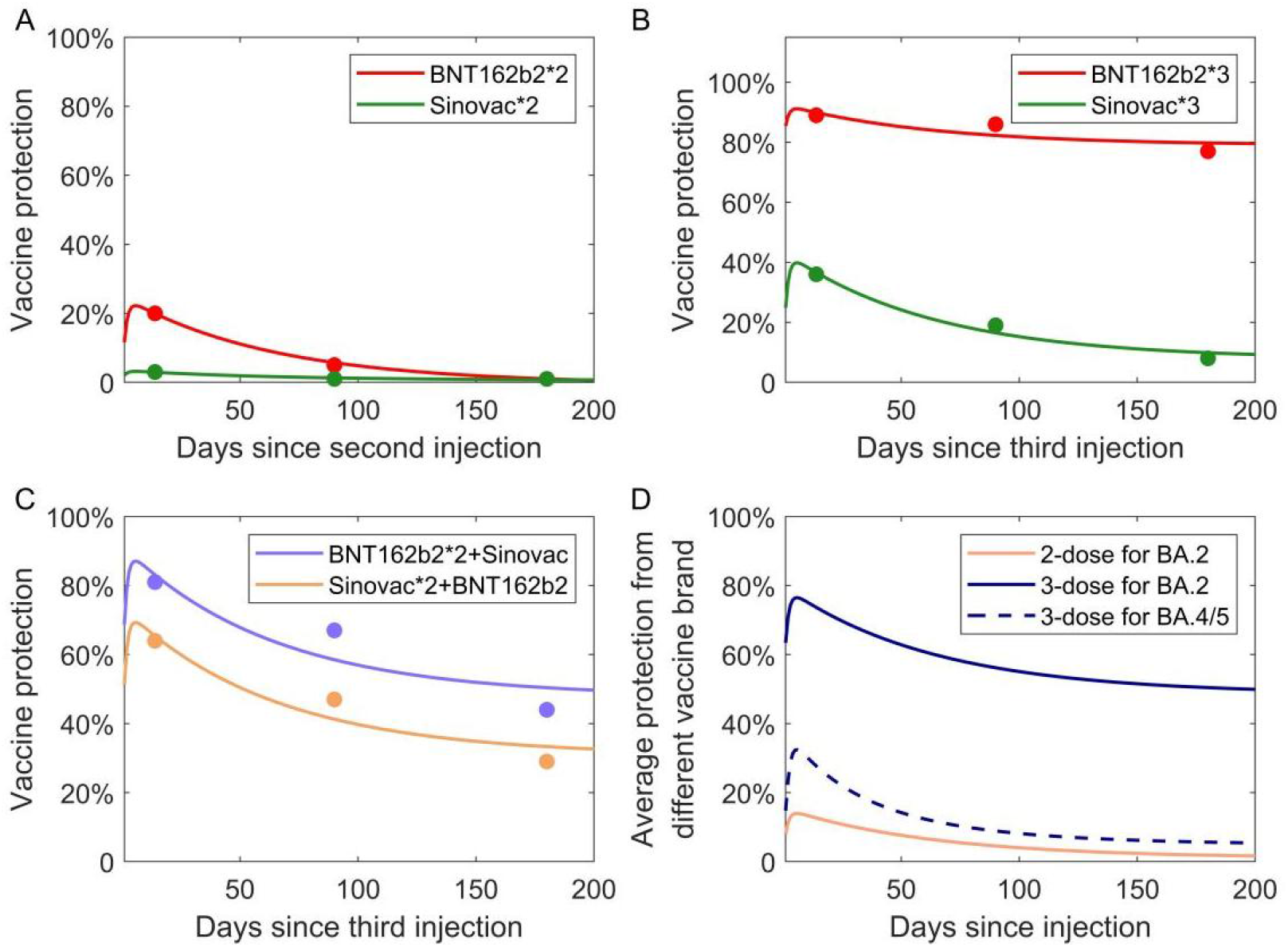
The simulation of waning immunity. (A) Vaccine protection from 2-dose. The red curve represents the protection of 2-dose Pfizer and the green curve represents the protection of 2-dose Sinovac. **(B-C) Vaccine protection from 3-dose**. The four curves represent the protection since the third dose with different vaccination schemes. **(D) Average vaccine protection from different vaccine brands and 3-dose protection for variant BA.4/5**. The pink curve is obtained from the arithmetic mean of the two curves in the A diagram. The blue curve is obtained from the arithmetic mean of the four curves in the **B** diagram. The dashed line indicates the protection of 3-dose for BA.4/5.

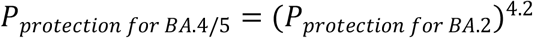

Moreover, the protection of one or two doses against BA.4/5 is assumed to be 0.

For illustrative purposes, we start with a model of two co-circulating strains within a simplified SEIR framework, assuming a complete cross-immunity. This means infection by either strain confers permanent immunity (at least in this model’s short-term period) to both. The distinct compartmental classes were: susceptible to both strains S, vaccinated group V, exposed to BA.2, exposed to BA.4/5, infectious with BA.2, infectious with BA.4/5, and recovered (immune to both).

### Model training and forecast

To predict the coming outbreak, we first fitted the fifth wave. 22nd February 2021 was the start of the vaccine and also was day 0 for model simulation. The parameterized model (**Supplementary Table S2**) was used to obtain the future daily number of reported cases and daily cases among individuals with a different number of vaccine doses.

The population is classified into different vaccination groups. We assumed that during the fifth epidemic wave, 95% of susceptible people who should have received the first dose (i.e. individuals who are eligible for the second injection) received the second dose, and only 70% of people who should have received the second dose received the third dose. After 5th March 2022, the average interval between the 2nd and 3rd dose was shortened to 90 days for 90% population (who would receive the second dose after 5th March 2022) while that interval for another 10% population remained unchanged (still 200 days). Next, the impacts of the new VOC on vaccine protection were incorporated into the modeling. Some assumptions were made: First, BA 2.12.1 and BA.2 were grouped because of their similar growth pattern in Hong Kong (see supplementary). Second, BA 4 and BA. 5 were not separated in the model [22]. Finally, there was complete cross-immunity between these two types, which meant that recovered individuals with either virus lead to immunity to both.

We projected outbreak dynamics by different social distancing measures and vaccination coverage rates. Three possible measures were proposed: (1) Reactivate the social distance policy [25]: the mobility index was decreased by 10% from 1^st^ September to 1^st^ October compared with the original forecast data; (2) Higher vaccinated coverage with boosting dose [26]: It was assumed that 100% people who never infected would finally complete three doses, to explore whether a larger scale of vaccine coverage combined with the recovered group could control the spread of new variants. (3) The above two measures were implemented simultaneously.

## Results

### The vaccine protection taking account of waning immunity

The detailed results of simulating vaccine waning immunity were presented in **Fig.1** The protections of full immunization (two doses) from each of two brands, homologous boosters and heterologous boosters were obtained. Then, we calculated the average protection provided from two doses against BA.2 and BA.4/5 from the actual vaccination rates and the protection from three doses. For simplicity purpose, only the average protection was used for simulation in this study (**Fig. 3C**).

Let 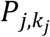 be the protection of the *j*-th dose over time. Define *V*^*max*^ as the maximum value of protection and *V*^*inf*^ as the minimum value of protection after waning. The protections of different doses over time are defined as:
1-dose: 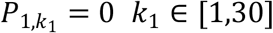

2- and 3-dose:

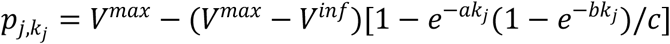

where the index *k*_*j*_ represents the time (number of days) since the j-th dose was administered. We set the range for *k*_1_, *k*_2_ and *k*_*3*_: *k*_1_ ∈ [1,*3*0] and *k*_*j*_ ∈ [1,200] when *j* = 2,*3*, considering the actual dose intervals recommended in Hong Kong. The parameters *a, b, c* are parameters to control the slopes of the functions.

### The fifth epidemic wave in Hong Kong

The model successfully predicted the evolving trend of the COVID-19 epidemic after the training period **(Fig. 4C)**. During the forecast period, BA.4/5 emerged and vaccine protection waned. Temperature generally became warmer while social distancing was relaxed (See Methods and **Supplementary Table S3**). The model was also able to predict daily injections of the second and third doses using the real vaccination data of the first dose after required dose intervals were taken into account **(Fig. 4AB)**.

**Fig. 4.**
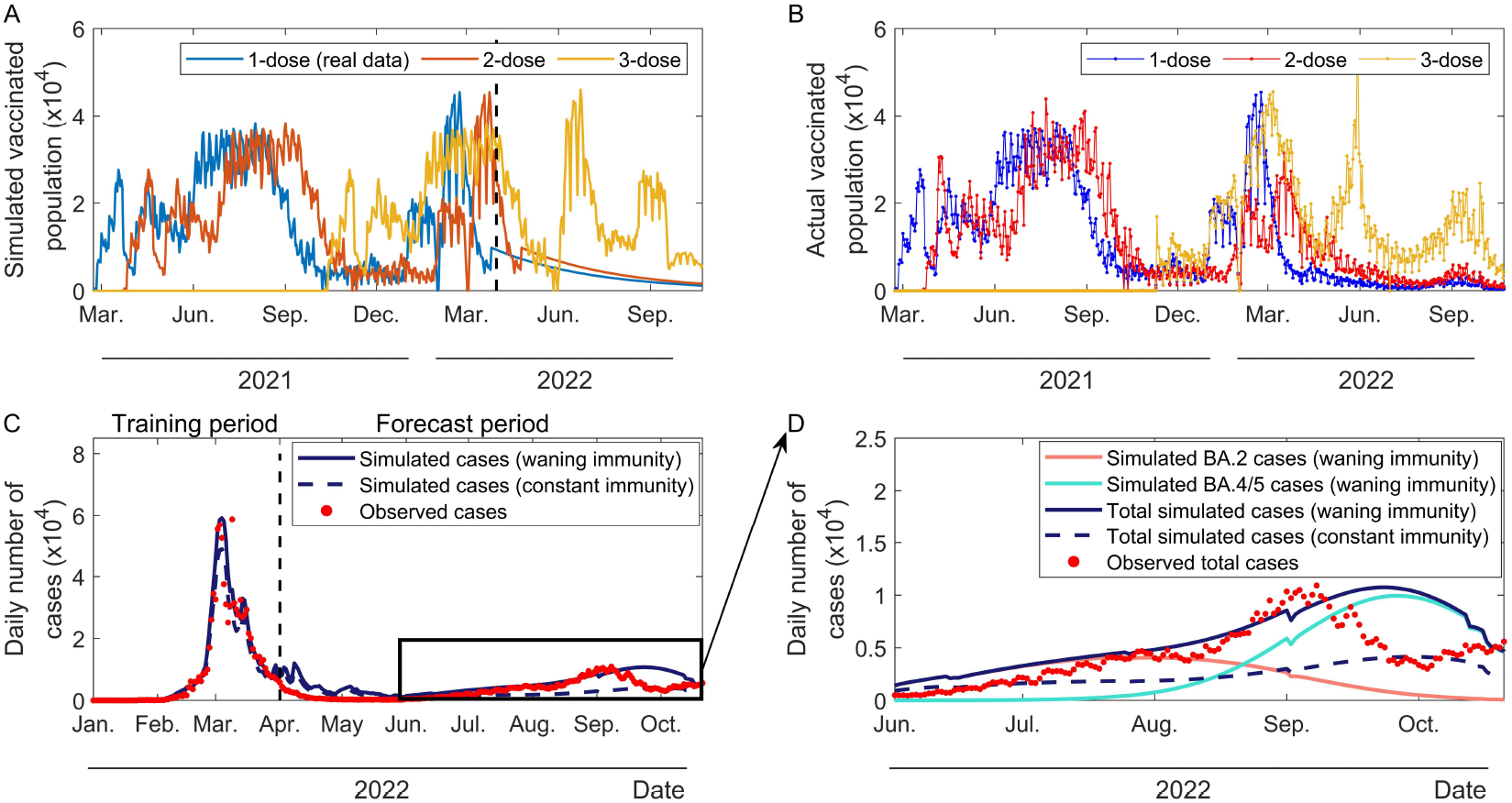
Overview of fifth epidemic curve estimation, predicted and actual daily number of vaccination. (A) Predicted ideal daily number of vaccination. (B) Actual daily number of vaccination(As of October 30th, 2022). (C) Simulated fifth epidemic curve and possible sixth wave with waning vaccine immunity and constant vaccine immunity. The red dots represent the observed daily reported cases. The dark blue curve represents the fitting daily number of infections with waning immunity. The dashed dark blue curve represents the fitting daily number of infections without waning immunity. **(D) The predicted number of daily infections with two variants’ (BA.2 and BA.4/5) cross-immunity with waning vaccine immunity and constant vaccine immunity**.

### Impacts of vaccine waning and immune escape

To verify the impact of waning immunity, a null model without waning immunity was compared. The average protection of 3rd dose was maintained at the maximum value without further decline (i.e. 76.48%). The null model cannot capture the actual transmission dynamics. The predicted number of cumulative cases across the study period was 35.14% fewer than the model with waning immunity **(Fig. 4CD)**.

The model forecasted the dynamics of multiple strains. BA.4/5 were assumed to enter the community on 1st June. The model predicted that BA.2 would cause an outbreak and reach its peak around mid-September. BA.4/5 infection would become the dominant strain after late August **(Fig. 5A)**. The infectious rate would become higher due to closer social distance, waning vaccine protection, and decreasing temperature in autumn and winter or other reasons (see method). These factors determined a new outbreak.

**Fig. 5.**
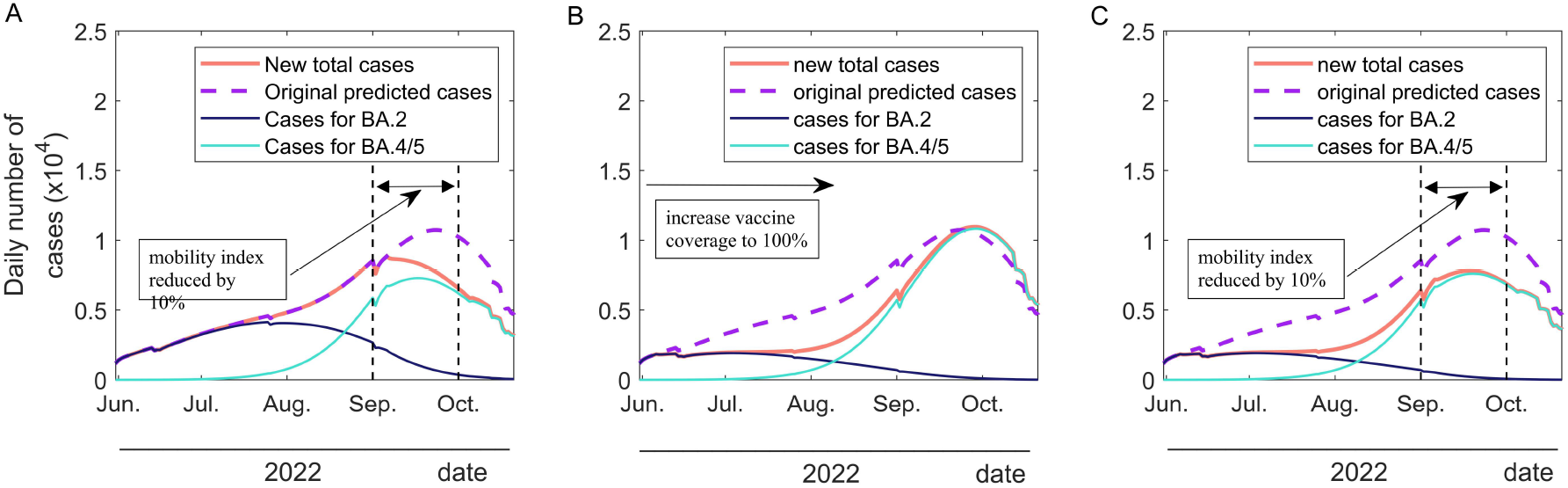
Number of predicted infections during second surge of Hong Kong’s fifth epidemic wave under variant BA.2 and BA.4/5. (A) The government re-publish social distance policy. The vertical dashed lines represent the period (between 1st September and 1st October) when the mobility index is reduced by 10%. (B) Fully vaccinated for boosting dose (after 1st June) The model assumed population would be fully vaccinated for boosting dose. (C) The policy that more stringent social distance and population would be fully vaccinated would be implemented together. The vertical dashed lines have the same meaning as in (A).

### Impacts of control measures

We projected the second surge under different social distancing and vaccination conditions. When the population mobility was reduced by 10% between 1st September 2022 and 1st October 2022, compared with the previous forecast data), the number of cases reduced 13.78% (about 121400). **(Fig. 5A)**.

If the probability of receiving boosting dose improved to 100% from 70% after 1st, June 2022, the BA.2 outbreak would be suppressed obviously. Although a higher vaccination coverage allows the population less likely to be infected with BA.2 and also BA.4/5, vaccines against BA.4/5 could not reduce the number of infections. 55.93 % of BA.2 cases can be reduced but cases of BA.4/5 are almost unchanged. Total cases would reduce 18.65% (**Fig. 5B**). If both measures were used, social distance policy and higher vaccination coverage could control possible next surge better and 33.39 % of total cases can be reduced (**Fig. 5C**). However, the pattern of the second surge remained apparently.

## Discussion

This study successfully forecasted the development of COVID-19’s second surge after the initial fifth epidemic wave in Hong Kong by incorporating a fine-scaled vaccine-waning immunity into an SEIR model. The results demonstrate that the waning protection and immune escape mutations were the most critical factors to drive this new outbreak. The impact of our finding is that waning immunity should be considered in order to provide a more accurate model forecast. A more accurate model forecasting considering waning immunity allows a better preparation of the next outbreaks. Incorporating the continuous change of vaccine protection into modelling can also be used to improve previous vaccine dose interval studies [9,10], aiming to optimize vaccine protection.

The comparison of the predictions between models with waning and constant immunity settings suggested that waning vaccine immunity is a factor leading to the second surge of infections **(Figure 5D)**. Moreover, around the second surge, two types of subvariants (BA.2 and BA.4/5) in the population co-circulated and produced sufficient cross-immunity against each other. There, we assumed that individuals were only infected with one of these two subvariants and then develop strong protection against the other at least during the study period (i.e. between January 2022 and November 2022). The model also successfully captured that after the two variants spread for a period, subvariant BA.4/5 gradually became the dominant strain. The replacement happened because vaccines only effectively protect BA.2 but not BA.4/5 (**Fig. 3D**). Even if vaccine coverage reached 100%, the surge dominated by BA.4/5 can not be prevented (**Fig. 5B**). This suggests BA4.5 had a stronger vaccine escape ability.

Besides the dynamics of population immunity and immune escape mutations, other factors such as temperature, contact number, and population mobility index was incorporated into modeling to capture the development of the fifth epidemic wave in Hong Kong. Although the temperature was likely to influence virus transmission significantly during the early period of the fifth wave [16], it was not the critical factor to lead to the second surge, which occurred before winter. Similarly, when the mobility index was further reduced, the second surge was maintained, indicating the new outbreaks caused by Omicron immune escape strains may not easily be prevented by non-pharmaceutical interventions.

Even though we successfully predicted the second surge, the number of reported cases reduced more rapidly than our predicted decline. Certain reasons may lead to this discrepancy. First, some continuous-executing policies such as the “Vaccine Pass” or gathering ban policy would help control the epidemic. “Vaccine Pass” required citizens to receive three doses of the COVID-19 vaccine as soon as possible (From 30^th^ June, 2022, population aged 18 and over must continue to receive the third dose of vaccine within 9 months of receiving the second dose of vaccine in order to continue to use the “Vaccine Pass”). The mobility index we used cannot reflect the impact of this policy. Second, many recovered people still get three doses of the vaccine or even a fourth dose. A high-level vaccine coverage would provide higher protection at the social level. Finally, people would consciously take some measures to protect themselves when facing a possible new wave, such as not going to crowded places and reducing the use of transportation, etc.

Certain limitations exist in our study. First, we assumed that the recovered population had long-term immunity. We acknowledged that recovered people still have a chance of being re-infected[27, 28], however, this event has not been frequently reported during this period. Second, the vaccine was assumed to be equally protective across different age groups and the duration of recovery for these age groups was only related to their vaccination status. Certain immunocompromised people (like the elderly, pregnant women, and children) can be more susceptible to COVID-19 infectious but we did not consider this scenario. Third, the model did not account for the fourth dose although it has entered the community, which was still significant to obtain a higher vaccine protection across the population [29,30]. Fourth, the model did not consider unreported cases. Since rapid antigen test kits were allowed to be used for the case confirmation in this period, the fraction of unreported might not be significant. Finally, only two types of subvariants (BA.2 and BA.4/5) were studied. There could be other strains circulated but we did not find the report of these events during most of study period.

## Data Availability

All data produced in the present study are available upon reasonable request to the authors

## Reference

1. https://www.who.int/publications/m/item/covid-19-public-health-emergency-of-international-concern-(pheic)-global-research-and-innovation-forum#:∼:text=On%2030%20January%202020%20following,of%20International%20Concern%20(PHEIC)

2. Levin EG, Lustig Y, Cohen C, et al. Waning Immune Humoral Response to BNT162b2 COVID-19 Vaccine over 6 Months. N Engl J Med. 2021;385(24):e84. doi:10.1056/NEJMoa2114583

3. Thompson MG, Burgess JL, Naleway AL, et al. Interim Estimates of Vaccine Effectiveness of BNT162b2 and mRNA-1273 COVID-19 Vaccines in Preventing SARS-CoV-2 Infection Among Health Care Personnel, First Responders, and Other Essential and Frontline Workers - Eight U.S. Locations, December 2020-March 2021. MMWR Morb Mortal Wkly Rep. 2021;70(13):495–500. Published 2021 Apr 2. doi:10.15585/mmwr.mm7013e3

4. Peng Q, Zhou R, Wang Y, et al. Waning immune responses against SARS-CoV-2 variants of concern among vaccinees in Hong Kong. EBioMedicine. 2022;77:103904. doi:10.1016/j.ebiom.2022.103904

5. Yuan HY, Liang J, Hossain MP. Impacts of social distancing, rapid antigen test and vaccination on the Omicron outbreak during large temperature variations in Hong Kong: A modelling study. J Infect Public Health. 2022;15(12):1427–1435. doi:10.1016/j.jiph.2022.10.0266.

6. https://www.info.gov.hk/gia/general/202211/01/P2022110100516.htm

7. Wang Q, Guo Y, Iketani S, et al. Antibody evasion by SARS-CoV-2 Omicron subvariants BA.2.12.1, BA.4 and BA.5. Nature. 2022;608(7923):603–608. doi:10.1038/s41586-022-05053-w

8. https://www.med.hku.hk/en/news/press//-/media/HKU-Med-Fac/News/slides/20220314-sims_wave_5_omicron_2022_03_14_final.ashx

9. Liu Y, Pearson CAB, Sandmann FG, et al. Dosing interval strategies for two-dose COVID-19 vaccination in 13 middle-income countries of Europe: Health impact modelling and benefit-risk analysis. Lancet Reg Health Eur. 2022;17:100381. doi:10.1016/j.lanepe.2022.100381

10. Bubar KM, Reinholt K, Kissler SM, et al. Model-informed COVID-19 vaccine prioritization strategies by age and serostatus. Science. 2021;371(6532):916–921. doi:10.1126/science.abe6959

11. Khoury, D. S., Cromer, D., Reynaldi, A., Schlub, T. E., Wheatley, A. K., Juno, J. A., Subbarao, K., Kent, S. J., Triccas, J. A., & Davenport, M. P. (2021). Neutralizing antibody levels are highly predictive of immune protection from symptomatic SARS-CoV-2 infection. Nature medicine, 27(7), 1205–1211. https://doi.org/10.1038/s41591-021-01377-8

12. He S, Peng Y, Sun K. SEIR modeling of the COVID-19 and its dynamics. Nonlinear Dyn. 2020;101(3):1667–1680. doi:10.1007/s11071-020-05743-y

13. Pérez-Then, E., Lucas, C., Monteiro, V. S., Miric, M., Brache, V., Cochon, L., Vogels, C., Malik, A. A., De la Cruz, E., Jorge, A., De Los Santos, M., Leon, P., Breban, M. I., Billig, K., Yildirim, I., Pearson, C., Downing, R., Gagnon, E., Muyombwe, A., Razeq, J., … Iwasaki, A. (2022). Neutralizing antibodies against the SARS-CoV-2 Delta and Omicron variants following heterologous CoronaVac plus BNT162b2 booster vaccination. Nature medicine, 28(3), 481–485. https://doi.org/10.1038/s41591-022-01705-6

14. Liang JB, Yuan HY, Li KK, et al. Path to normality: Assessing the level of social-distancing measures relaxation against antibody-resistant SARS-CoV-2 variants in a partially-vaccinated population. Comput Struct Biotechnol J. 2022;20:4052–4059. doi:10.1016/j.csbj.2022.07.048

15. https://ourworldindata.org/covid-google-mobility-trends

16. Sera F, Armstrong B, Abbott S, et al. A cross-sectional analysis of meteorological factors and SARS-CoV-2 transmission in 409 cities across 26 countries. Nat Commun. 2021;12(1):5968. Published 2021 Oct 13. doi:10.1038/s41467-021-25914-8

17. Ahmed I, Modu GU, Yusuf A, Kumam P, Yusuf I. A mathematical model of Coronavirus Disease (COVID-19) containing asymptomatic and symptomatic classes. Results Phys. 2021;21:103776. doi:10.1016/j.rinp.2020.103776.

18. Childs L, Dick DW, Feng Z, Heffernan JM, Li J, Röst G. Modeling waning and boosting of COVID-19 in Canada with vaccination. Epidemics. 2022;39:100583. doi:10.1016/j.epidem.2022.100583

19. Dick DW, Childs L, Feng Z, et al. COVID-19 Seroprevalence in Canada Modelling Waning and Boosting COVID-19 Immunity in Canada a Canadian Immunization Research Network Study. Vaccines (Basel). 2021;10(1):17. Published 2021 Dec 23. doi:10.3390/vaccines10010017

20. Diagne ML, Rwezaura H, Tchoumi SY, Tchuenche JM. A Mathematical Model of COVID-19 with Vaccination and Treatment. Comput Math Methods Med. 2021;2021:1250129. Published 2021 Sep 4. doi:10.1155/2021/1250129

21. Azimi P, Keshavarz Z, Cedeno Laurent JG, Stephens B, Allen JG. Mechanistic transmission modeling of COVID-19 on the Diamond Princess cruise ship demonstrates the importance of aerosol transmission. Proc Natl Acad Sci U S A. 2021;118(8):e2015482118. doi:10.1073/pnas.2015482118

22. Qu P, Faraone JN, Evans JP, et al. Durability of the Neutralizing Antibody Response to mRNA Booster Vaccination Against SARS-CoV-2 BA.2.12.1 and BA.4/5 Variants. Preprint. bioRxiv. 2022;2022.07.21.501010. Published 2022 Jul 22. doi:10.1101/2022.07.21.50101

23. Thompson MG, Burgess JL, Naleway AL, et al. Prevention and Attenuation of Covid-19 with the BNT162b2 and mRNA-1273 Vaccines. N Engl J Med. 2021;385(4):320–329. doi:10.1056/NEJMoa2107058

24. Thompson MG, Burgess JL, Naleway AL, et al. Prevention and Attenuation of COVID-19 with the BNT162b2 and mRNA-1273 Vaccines. N Engl J Med. 2021;385(4):320–329. doi:10.1056/NEJMoa2107058

25. Mwalili S, Kimathi M, Ojiambo V, Gathungu D, Mbogo R. SEIR model for COVID-19 dynamics incorporating the environment and social distancing. BMC Res Notes. 2020;13(1):352. Published 2020 Jul 23. doi:10.1186/s13104-020-05192-1

26. Christie A, Brooks JT, Hicks LA, et al. Guidance for Implementing COVID-19 Prevention Strategies in the Context of Varying Community Transmission Levels and Vaccination Coverage. MMWR Morb Mortal Wkly Rep. 2021;70(30):1044–1047. Published 2021 Jul 27. doi:10.15585/mmwr.mm7030e2

27. Kojima N, Klausner JD. Protective immunity after recovery from SARS-CoV-2 infection. Lancet Infect Dis. 2022;22(1):12–14. doi:10.1016/S1473-3099(21)00676-9

28. Kwok KO, Li KK, Tang A, et al. Psychobehavioral Responses and Likelihood of Receiving COVID-19 Vaccines during the Pandemic, Hong Kong. Emerg Infect Dis. 2021;27(7):1802–1810. doi:10.3201/eid2707.210054

29. Bar-On YM, Goldberg Y, Mandel M, et al. Protection by a Fourth Dose of BNT162b2 against Omicron in Israel. N Engl J Med. 2022;386(18):1712–1720. doi:10.1056/NEJMoa2201570

30. Gazit S, Saciuk Y, Perez G, Peretz A, Pitzer VE, Patalon T. Short term, relative effectiveness of four doses versus three doses of BNT162b2 vaccine in people aged 60 years and older in Israel: retrospective, test negative, case-control study. BMJ. 2022;377:e071113. Published 2022 May 24. doi:10.1136/bmj-2022-071113

