## Supplementary for "Vaccine waning and immune escape drive the second surge of Omicron spread in Hong Kong: A modeling study"

### Supplementary material

#### Simulation for waning immunity from vaccine

##### *The proportion of different vaccination schemes*

As of June 30, 2022, Hong Kong residents have received a total of 6,454,900 second doses vaccine: BNT162b2(3655554 doses, 56.6%) and SINOVAC(2799346, 43.4%)[1]. Assume that the second dose vaccine must be the same brand as the first dose. The proportions of the two vaccination schemes in **Fig. 3A** were 56.6% and 43.3% from top to bottom, respectively. Similarly, As of June 30, 2022, Hong Kong residents have received a total of 4360552 third doses vaccine: BNT162b2(2866282 doses, 65.7%) and SINOVAC(1494260, 34.3%)[1]. The proportions of the four vaccination schemes in **Fig. 3B** are:

$$\text{BNT162b2*3: } 56.6\% \times 65.7\% = \mathbf{37.19\%}$$

$$\text{BNT162b2*2+SINOVAC: } 56.6\% \times 34.3\% = \mathbf{19.41\%}$$

$$\text{SINOVAC*2+BNT162b2: } 43.4\% \times 65.7\% = \mathbf{28.51\%}$$

$$\text{SINOVAC*3: } 43.4\% \times 34.3\% = \mathbf{14.89\%}$$

##### *Data for vaccine's protectiveness*

| VE in reducing susceptibility | | Time since 2nd or 3rd dose | | |
|---|---|---|---|---|
| Virus | Vaccine | 14 days | 90days | 180days |
| Omicron 1 dose | BNT162b2*1 | 0 | 0 | 0 |
| | Sinovac*1 | 0 | 0 | 0 |
| Omicron 2 dose | BNT162b2*2 | 0.20 | 0.05 | 0.01 |
| | Sinovac*2 | 0.03 | 0.01 | 0.01 |
| Omicron 3 dose | BNT162b2*3 | 0.89 | 0.86 | 0.77 |
| | BNT162b2*2+Sinovac | 0.81 | 0.67 | 0.44 |
| | Sinovac*2+BNT162b2 | 0.64 | 0.47 | 0.29 |
| | Sinovac*3 | 0.36 | 0.19 | 0.08 |

**Table S1. Estimates of vaccine effectiveness in reducing susceptibility by time since the second or third dose [2]**

#### Mathematical expression of discrete-time Vaccinated SEIR model

A discrete-time model with vaccination was developed based on SEIR framework. The model further subdivides the vaccinated population into subgroups according to the number of doses and the number of days since vaccination. This part presents the Detailed description and assumption in Method **Fig.1** and **Fig.2**.

##### *Detailed SEIR flow*

Vaccinated individuals were classified into 3 statuses ($V_1, V_2$ and $V_3$) depending on the number of doses received. The vaccine protections are changed every day. We subdivided *V* according to the number of days after vaccinate ($V_{1,1}, V_{1,2}...V_{1,30}$; $V_{2,1}, V_{2,2}...V_{2,200}$ and $V_{3,1}, V_{3,2}...V_{3,200}...$). After being infected but not yet being infectious, parts of susceptible individuals (*S*) and vaccinated individuals (*V*) become exposed (*E*) every day different virus daily transmission rate $\beta_{j,k}$, which is changed by the number of vaccines already administered, the number of days after the last dose, daily average mobility index, vaccine protection and daily temperature. Exposed individuals became infectious individuals (*I*) after a latent period ($1/\delta$). Similar to $V_{j,k}$, we obtain $E_{j,k}$ ($j$ is the number of vaccines already administered and $k$ is the number of days after the last dose). Finally, infectious individuals recovered with recovered rate ($\gamma$) and became recovered group (*R*). $\gamma_0$ is the recovered rate of individuals without vaccination and $\gamma_v$ is the recovered rate of individuals with vaccination.

There were key model assumptions: First, all individuals in *E* group would finally become *I* group. Second, people who recovered from infection had long-term immunity (at least within half or one year) and their protections for any COVID-19 virus are 100%. Third, all eligible people received the first dose. People who have received the first dose and never been infected would received the second dose with the probability $pro_2$. Similarly, People who had received the second dose and never been infected would received the third dose with the probability $pro_3$. The vaccine protection of people who did not received the next dose declined to 0. Fourth, all individuals in *E* group will finally become I group. In other words, if someone gets ill, he will be found, not before he heals. Last, asymptomatic infections were not combined in cases. The details of the model were summarized in **Fig.S1**.

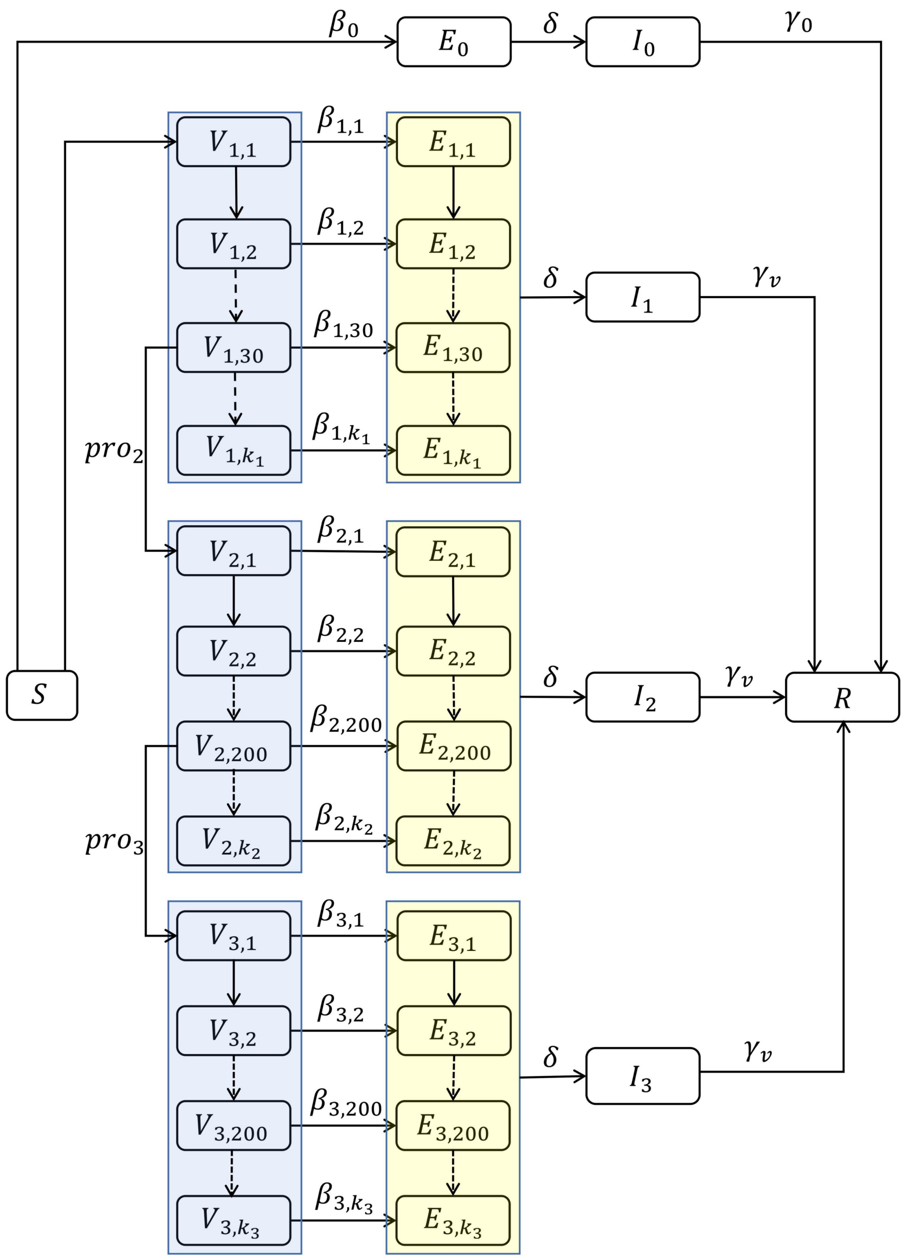


**Fig. S1. Daily time-step discrete SEIR model with vaccination.** Individuals were classified into the following group: *S* (Susceptible, without any vaccination and never been infected), *V* (Individuals with vaccination), *E* (exposed but not yet infected individuals), *I* (infected individuals, including pre-clinical, clinical, and sub-clinical infections) and *R* (recovered, including hospital confirmed, recovered and removed individuals).

#### *Mathematical expression*

The susceptible population (without vaccine) at the end of the *t*-th day is represented by $\boldsymbol{S^t}$; The exposed population (have been infected but not infectious) at the end of the *t*-th day is represented by $\boldsymbol{E^t}$; The infectious population at the end of the *t*-th day is represented by $\boldsymbol{I^t}$ . The dynamics of infectious population without vaccination is described as follows:

$$I_{total}^{t-1} = I_0^{t-1} + \sum_{k_1=1}^{d_1} I_{1,k_1}^{t-1} + \sum_{k_2=1}^{d_2} I_{2,k_2}^{t-1} + \sum_{k_3=1}^{d_3} I_{3,k_3}^{t-1}$$

$$S^t = S^{t-1} - \frac{1}{N} S^{t-1} \beta_0^{t-1} I_{total}^{t-1} \Delta T - v_1^{t-1}$$

$$E_0^t = \frac{1}{N} S^{t-1} \beta_0^{t-1} I_{total}^{t-1} \Delta T + (1-\delta) E_0^{t-1}$$

$$I_0^t = \delta E_0^{t-1} + (1-\gamma_0) I_0^{t-1}$$

where $d_1$ is the maximum period between the first and the second vaccine; $d_2$ is the maximum period between the second and the third vaccine; $d_3$ is the maximum period between the third vaccine and the time of completion of the study; $\gamma_0$ is the recovered rate of individuals without vaccination; δ is transport rate from *E* group into *I* group; $\beta$ is the ability of disease transmission across the population. $\Delta T$ is time-step.

Next, we define the notations for vaccinated groups: $\boldsymbol{V_{j,k}^t}$ is the vaccinated population who received the *j*-th dose vaccine *k* days ago at the end of the *t*-th day. $\boldsymbol{E_{j,k}^t}$ is the exposed population who were infected on the *k*-th day after receiving *j*-th dose vaccine. Similar, infectious population is $\boldsymbol{I_{j,k}^t}$. The dynamics of infectious populations with vaccination are described as follows:

1. For people who have received one dose:

$V_{1,1}^t = v_1^{t-1}$

$V_{1,k_1}^t = V_{1,k_1-1}^{t-1} - \frac{1}{N} V_{1,k_1-1}^{t-1} \beta_{1,k_1-1}^{t-1} I_{total}^{t-1} \Delta T - pro_2 \cdot v_{2,k_1-1}^{t-1}$ $\quad (k_1 = 2,3,\ldots\ldots d_1)$

$V_{1,k_1}^t = V_{1,k_1-1}^{t-1} - \frac{1}{N} V_{1,k_1-1}^{t-1} \beta_0^{t-1} I_{total}^{t-1} \Delta T$ $\quad (k_1 > d_1)$

$E_{1,k_1}^t = \frac{1}{N} V_{1,k_1}^{t-1} \beta_{1,k_1}^{t-1} I_{total}^{t-1} \Delta T - (1-\delta) E_{1,k_1}^{t-1}$ $\quad (k_1 = 1,2,3,\ldots\ldots d_1)$

$E_{1,k_1}^t = \frac{1}{N} V_{1,k_1}^{t-1} \beta_0^{t-1} I_{total}^{t-1} \Delta T - (1-\delta) E_{1,k_1}^{t-1}$ $\quad (k_1 > d_1)$

$I_{1,k_1}^t = (1-\gamma_v) I_{1,k_1}^{t-1} + \delta E_{1,k_1}^{t-1}$

2. For people who have received two doses:

$$V_{2,1}^{t} = \sum_{k_1=1}^{d_1} pro_2 \cdot v_{2,k_1}^{t-1}$$

$$V_{2,k_2}^{t} = V_{2,k_2-1}^{t-1} - \frac{1}{N} V_{2,k_2-1}^{t-1} \beta_{2,k_2-1}^{t-1} I_{total}^{t-1} \Delta T - pro_3 \cdot v_{3,k_2-1}^{t-1} \qquad (k_2 = 2,3,\ldots\ldots d_2)$$

$$V_{2,k_2}^{t} = V_{2,k_2-1}^{t-1} - \frac{1}{N} V_{2,k_2-1}^{t-1} \beta_0^{t-1} I_{total}^{t-1} \Delta T \qquad (k_2 > d_2)$$

$$E_{2,k_2}^{t} = \frac{1}{N} V_{2,k_2}^{t-1} \beta_{2,k_2}^{t-1} I_{total}^{t-1} \Delta T - (1-\delta) E_{2,k_2}^{t-1} \qquad (k_2 = 1,2,3,\ldots\ldots d_2)$$

$$E_{2,k_2}^{t} = \frac{1}{N} V_{2,k_2}^{t-1} \beta_0^{t-1} I_{total}^{t-1} \Delta T - (1-\delta) E_{2,k_2}^{t-1} \qquad (k_2 > d_2)$$

$$I_{2,k_2}^{t} = (1-\gamma_v) I_{2,k_2}^{t-1} + \delta E_{2,k_2}^{t-1}$$

3. For people who have received three doses:

$$V_{3,1}^{t} = \sum_{k_2=1}^{d_2} pro_3 \cdot v_{3,k_2}^{t-1}$$

$$V_{3,k_3}^{t} = V_{3,k_3-1}^{t-1} - \frac{1}{N} V_{3,k_3-1}^{t-1} \beta_{3,k_3-1}^{t-1} I_{total}^{t-1} \Delta T \qquad (k_3 = 2,3,\ldots\ldots d_3)$$

$$V_{3,k_3}^{t} = V_{3,k_3-1}^{t-1} - \frac{1}{N} V_{3,k_3-1}^{t-1} \beta_0^{t-1} I_{total}^{t-1} \Delta T \qquad (k_3 > d_3)$$

$$E_{3,k_3}^{t} = \frac{1}{N} V_{3,k_3}^{t-1} \beta_{3,k_3}^{t-1} I_{total}^{t-1} \Delta T - (1-\delta) E_{3,k_3}^{t-1} \qquad (k_3 = 1,2,3,\ldots\ldots d_3)$$

$$E_{3,k_3}^{t} = \frac{1}{N} V_{3,k_3}^{t-1} \beta_0^{t-1} I_{total}^{t-1} \Delta T - (1-\delta) E_{3,k_3}^{t-1} \qquad (k_3 > d_3)$$

$$I_{3,k_3}^{t} = (1-\gamma_v) I_{3,k_3}^{t-1} + \delta E_{3,k_3}^{t-1}$$

where $v_1^t$ is the number of injections of the first-dose vaccine at the end of the *t*-th day ; $v_{2,k_2}^t$ ($v_{3,k_3}^t$) is the number of injections of the second (third)-dose vaccination at the end of the *t*-th day.

***Cross immunity model***

Similarly, we added another variant of concern BA.4/5 in following model based on the model above:

$$\beta_{B\_j,k}^{t} = C_0 \cdot M_t \cdot (1 - p_{j,k}) \cdot \rho_B \cdot W_t$$

$\rho_B$ is the probability of mutation BA.2's successful transmission under each contact with each individual;

Suppose:

$$I_{total}^{t-1} = I_0^{t-1} + \sum_{k_1=1}^{d_1} I_{1,k_1}^{t-1} + \sum_{k_2=1}^{d_2} I_{2,k_2}^{t-1} + \sum_{k_3=1}^{d_3} I_{3,k_3}^{t-1}$$

$$I_{B\_total}^{t-1} = I_{B\_0}^{t-1} + \sum_{k_1=1}^{d_1} I_{B\_1,k_1}^{t-1} + \sum_{k_2=1}^{d_2} I_{B\_2,k_2}^{t-1} + \sum_{k_3=1}^{d_3} I_{B\_3,k_3}^{t-1}$$

0-dose:

$$S^t = S^{t-1} - \frac{1}{N} S^{t-1} \beta_0^{t-1} I_{total}^{t-1} \Delta T - \frac{1}{N} S^{t-1} \beta_{0\_B}^{t-1} I_{B\_total}^{t-1} \Delta T - v_1^{t-1}$$

$$E_0^t = \frac{1}{N} S^{t-1} \beta_0^{t-1} I_{total}^{t-1} \Delta T + (1 - \delta) E_0^{t-1}$$

$$E_{B\_0}^t = \frac{1}{N} S^{t-1} \beta_{B\_0}^{t-1} I_{B\_total}^{t-1} \Delta T + (1 - \delta) E_{B\_0}^{t-1}$$

$$I_0^t = \delta E_0^{t-1} + (1 - \gamma_0) I_0^{t-1}$$

$$I_{B\_0}^t = \delta E_{B\_0}^{t-1} + (1 - \gamma_0) I_{B\_0}^{t-1}$$

1-dose:

$$V_{1,1}^t = v_1^{t-1}$$

$$V_{1,k_1}^t = V_{1,k_1-1}^{t-1} - \frac{1}{N} V_{1,k_1-1}^{t-1} \beta_{1,k_1-1}^{t-1} I_{total}^{t-1} \Delta T - \frac{1}{N} V_{1,k_1-1}^{t-1} \beta_{B\_1,k_1-1}^{t-1} I_{B\_total}^{t-1} \Delta T - pro_2 \cdot v_{2,k_1-1}^{t-1}$$

$$(k_1 = 2,3,\ldots\ldots d_1)$$

$$V_{1,k_1}^t = V_{1,k_1-1}^{t-1} - \frac{1}{N} V_{1,k_1-1}^{t-1} \beta_0^{t-1} I_{total}^{t-1} \Delta T - \frac{1}{N} V_{1,k_1-1}^{t-1} \beta_{B\_0}^{t-1} I_{B\_total}^{t-1} \Delta T \qquad (k_1 > d_1)$$

$$E_{1,k_1}^t = \frac{1}{N} V_{1,k_1}^{t-1} \beta_{1,k_1}^{t-1} I_{total}^{t-1} \Delta T - (1 - \delta) E_{1,k_1}^{t-1} \qquad (k_1 = 1,2,3,\ldots\ldots d_1)$$

$$E_{1,k_1}^{t} = \frac{1}{N} V_{1,k_1}^{t-1} \beta_0^{t-1} I_{total}^{t-1} \Delta T - (1-\delta) E_{1,k_1}^{t-1} \qquad (k_1 > d_1)$$

$$E_{B\_1,k_1}^{t} = \frac{1}{N} V_{1,k_1}^{t-1} \beta_{B\_1,k_1}^{t-1} I_{B\_total}^{t-1} \Delta T - (1-\delta) E_{B\_1,k_1}^{t-1} \qquad (k_1 = 1,2,3, \ldots\ldots d_1)$$

$$E_{B\_1,k_1}^{t} = \frac{1}{N} V_{1,k_1}^{t-1} \beta_{B\_0}^{t-1} I_{B\_total}^{t-1} \Delta T - (1-\delta) E_{B\_0,k_1}^{t-1} \qquad (k_1 > d_1)$$

$$I_{1,k_1}^{t} = (1-\gamma_v) I_{1,k_1}^{t-1} + \delta E_{1,k_1}^{t-1}$$

$$I_{B\_1,k_1}^{t} = (1-\gamma_v) I_{B\_1,k_1}^{t-1} + \delta E_{B\_1,k_1}^{t-1}$$

2-dose:

$$V_{2,1}^{t} = \sum_{k_1=1}^{d_1} pro_2 \cdot v_{2,k_1}^{t-1}$$

$$V_{2,k_2}^{t} = V_{2,k_2-1}^{t-1} - \frac{1}{N} V_{2,k_2-1}^{t-1} \beta_{2,k_2-1}^{t-1} I_{total}^{t-1} \Delta T - \frac{1}{N} V_{2,k_2-1}^{t-1} \beta_{B\_2,k_2-1}^{t-1} I_{B\_total}^{t-1} \Delta T - pro_3 \cdot v_{3,k_2-1}^{t-1}$$

$$(k_1 = 2,3, \ldots\ldots d_2)$$

$$V_{2,k_2}^{t} = V_{2,k_2-1}^{t-1} - \frac{1}{N} V_{2,k_2-1}^{t-1} \beta_0^{t-1} I_{total}^{t-1} \Delta T - \frac{1}{N} V_{2,k_2-1}^{t-1} \beta_{B\_0}^{t-1} I_{B\_total}^{t-1} \Delta T \qquad (k_2 > d_2)$$

$$E_{2,k_2}^{t} = \frac{1}{N} V_{2,k_2}^{t-1} \beta_{2,k_2}^{t-1} I_{total}^{t-1} \Delta T - (1-\delta) E_{2,k_2}^{t-1} \qquad (k_2 = 1,2,3, \ldots\ldots d_2)$$

$$E_{2,k_2}^{t} = \frac{1}{N} V_{2,k_2}^{t-1} \beta_0^{t-1} I_{total}^{t-1} \Delta T - (1-\delta) E_{2,k_2}^{t-1} \qquad (k_2 > d_2)$$

$$E_{B\_2,k_2}^{t} = \frac{1}{N} V_{2,k_2}^{t-1} \beta_{B\_2,k_2}^{t-1} I_{B\_total}^{t-1} \Delta T - (1-\delta) E_{B\_2,k_2}^{t-1} \qquad (k_2 = 1,2,3, \ldots\ldots d_2)$$

$$E_{B\_2,k_2}^{t} = \frac{1}{N} V_{2,k_2}^{t-1} \beta_{B\_0}^{t-1} I_{B\_total}^{t-1} \Delta T - (1-\delta) E_{B\_0,k_2}^{t-1} \qquad (k_2 > d_2)$$

$$I_{2,k}^{t} = (1-\gamma_v) I_{2,k}^{t-1} + \delta E_{2,k}^{t-1}$$

$$I_{B\_2,k}^{t} = (1-\gamma_v) I_{B\_2,k}^{t-1} + \delta E_{B\_2,k}^{t-1}$$

3-dose

$$V_{3,1}^{t} = \sum_{k_2=1}^{d_2} pro_3 \cdot v_{3,k_2}^{t-1}$$

$$V_{3,k_3}^{t}=V_{3,k_3-1}^{t-1}-\frac{1}{N}V_{3,k_3-1}^{t-1}\beta_{3,k_3-1}^{t-1}I_{total}^{t-1}\Delta T-\frac{1}{N}V_{B\_3,k_3-1}^{t-1}\beta_{B\_3,k_3-1}^{t-1}I_{B\_total}^{t-1}\Delta T$$

$$(k_3=2,3,\ldots\ldots d_3)$$

$$V_{3,k_3}^{t}=V_{3,k_3-1}^{t-1}-\frac{1}{N}V_{3,k_3-1}^{t-1}\beta_0^{t-1}I_{total}^{t-1}\Delta T-\frac{1}{N}V_{B\_0}^{t-1}\beta_{B\_3,k_3-1}^{t-1}I_{B\_total}^{t-1}\Delta T \qquad (k_3>d_3)$$

$$E_{3,k_3}^{t}=\frac{1}{N}V_{3,k_3}^{t-1}\beta_{3,k_3}^{t-1}I_{total}^{t-1}\Delta T-(1-\delta)E_{3,k_3}^{t-1} \qquad (k_3=1,2,3,\ldots\ldots d_3)$$

$$E_{3,k_3}^{t}=\frac{1}{N}V_{3,k_3}^{t-1}\beta_0^{t-1}I_{total}^{t-1}\Delta T-(1-\delta)E_{3,k_3}^{t-1} \qquad (k_3>d_3)$$

$$E_{3,k_3}^{t}=\frac{1}{N}V_{3,k_3}^{t-1}\beta_{B\_3,k_3}^{t-1}I_{total}^{t-1}\Delta T-(1-\delta)E_{B\_3,k_3}^{t-1} \qquad (k_3=1,2,3,\ldots\ldots d_3)$$

$$E_{3,k_3}^{t}=\frac{1}{N}V_{3,k_3}^{t-1}\beta_0^{t-1}I_{total}^{t-1}\Delta T-(1-\delta)E_{B\_3,k_3}^{t-1} \qquad (k_3>d_3)$$

$$I_{3,k_3}^{t}=(1-\gamma_v)I_{3,k_3}^{t-1}+\delta E_{3,k_3}^{t-1}$$

$$I_{B\_3,k_3}^{t}=(1-\gamma_v)I_{B\_3,k_3}^{t-1}+\delta E_{B\_3,k_3}^{t-1}$$

For population without vaccine, $E_0^t$ was the exposed population who were infected by mutation BA.2 and the corresponding infectious population is $I_0^t$. $E_{B\_0}^t$ was the exposed population who were infected by mutation BA.4/5 and the corresponding infectious population is $I_{B\_0}^t$. $E_{B\_j,k}^t$ was the exposed population who were infected by mutation BA.4/5 on the *k*-th day after the *j*-th vaccination and the corresponding infectious population is $I_{B\_j,k}^t$. $\beta_B$ describe the transmission ability for mutation variant BA.4/5 and β describe the transmission ability for normal strain BA.2.

#### Parameters

| Parameters | Description | Values | Reference |
|---|---|---|---|
| $V_{2-\text{BNT162b2}}^{max}$ | Maximum waning protection for 2-dose BNT162b2 | 0.2222 | Estimated |
| $V_{2-\text{BNT162b2}}^{inf}$ | Limit value for waning immunity for 2-dose BNT162b2 | 0 | Ditto |
| $V_{2-\text{Sinovac}}^{max}$ | Maximum waning protection for 2-dose Sinovac | 0.0318 | Ditto |
| $V_{2-\text{Sinovac}}^{inf}$ | Limit value for waning immunity for 2-dose Sinovac | 0.0059 | Ditto |
| $V_{3-\text{BNT162b2}}^{max}$ | Maximum waning protection for 3-dose BNT162b2 | 0.9114 | Ditto |
| $V_{3-\text{BNT162b2}}^{inf}$ | Limit value for waning immunity for 3-dose BNT162b2 | 0.7897 | Ditto |
| $V_{2-\text{BNT+Sino}}^{max}$ | Maximum waning protection for 2-dose BNT162b2 plus Sinovac | 0.8706 | Ditto |
| $V_{2-\text{BNT+Sino}}^{inf}$ | Limit value for waning immunity for 2-dose BNT162b2 plus Sinovac | 0.4778 | Ditto |
| $V_{2-\text{Sino+BNT}}^{max}$ | Maximum waning protection for 2-dose Sinovac plus BNT162b2 | 0.6930 | Ditto |
| $V_{2-\text{Sino+BNT}}^{inf}$ | Limit value for waning immunity for 2-dose Sinovac plus BNT162b2 | 0.3855 | Ditto |
| $V_{3-\text{Sino}}^{max}$ | Maximum waning protection for 3-dose Sinovac | 0.3985 | Ditto |
| $V_{3-\text{Sino}}^{inf}$ | Limit value for waning immunity for 3-dose Sinovac | 0.0774 | Ditto |
| $V_{2}^{max}$ | Average maximum waning protection from different brands for 2-dose vaccine | 0.1396 | Ditto |
| $V_{2}^{inf}$ | Average limit value for waning immunity for 2-dose vaccine | 0.1296 | Ditto |
| $V_{3}^{max}$ | Average maximum waning | 0.7648 | Ditto |

| | | | |
|---|---|---|---|
| | protection from different brands for 3-dose vaccine | | |
| $V_3^{inf}$ | Average limit value for waning immunity for 3-dose vaccine | 0.4856 | Ditto |
| $a$ | Parameters which describe waning rate of vaccine protection | $-ln3/70$ | Ditto |
| $b$ | Ditto | $-ln100/7$ | Ditto |
| $c$ | Ditto | 0.8926 | Ditto |
| $N$ | Total population in Hong Kong | 7.48 million | [3] |
| δ | Transport rate of infectious group from exposed group; | $1/3$ | [4] |
| $\gamma_0$ | Recovery rate for people without vaccination; | $1/8.9$ | [5] |
| $\gamma_v$ | Recovery rate for people with vaccination; | $1/2.7$ | [5] |
| $v_1^t$ | Number of first-dose vaccine injection; | Before 26/03/22: Real data On and after 26/03/22: $0.01 * S^{t-1}$ | Real data and Assumption |
| $v_{2,k_2}^t$ | Number of second-dose vaccine injection ; | $V_{1,d_1}^t$when $k = d_1$; 0 when $i < d_1$ | Assumption |
| $v_{3,k_3}^t$ | Number third-dose vaccine injection; | $V_{2,d_2}^t$when $k = d_2$; 0 when $i < d_2$ | Assumption |
| $d_1$ | Maximum time period between the first and the second vaccination; | 30 days | Assumption |
| $d_2$ | Maximum time period between the second and the third vaccination; | 200 days | Assumption |
| $d_3$ | Maximum length of period a third dose vaccine was effective; | 200 days | Assumption |
| $C_0$ | Average contact number; | 16.876 | [6] |

| | | | |
|---|---|---|---|
| $\rho_0$ | The probability of mutation BA.2's successful transmission under each contact with each individual; | 0.0313412 | Estimated |
| $\rho_B$ | The probability of mutation BA.2's successful transmission under each contact with each individual; | 0.0421872 | Estimated |
| $M_t$ | Population mobility index | Time-varying data | [7] |
| $W_t$ | Transmission effector affected by weather ($T_t$ is average temperature on t day [8] ) | $W_t = 0.00215T_t^3 - 0.0929875T_t^2 + 0.977175T_t + 1$ | Estimated |
| $pro_2$ | The probability for an individual who never been infected to get the second dose of vaccine | 0.95 | Assumption |
| $pro_3$ | The probability for an individual who never been infected to get the third dose of vaccine | 0.7 | Assumption |

**Table S2. Some parameters' explanation and value**

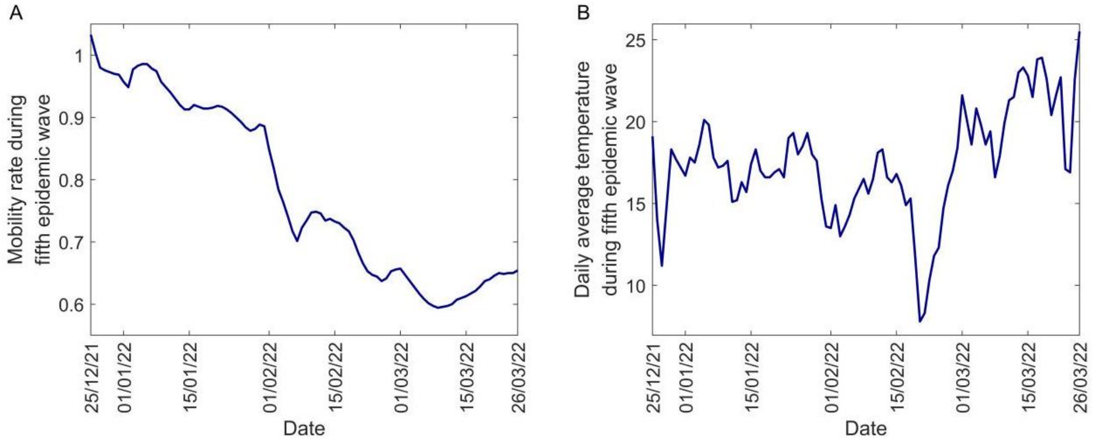


**Fig. S2. Diagram of parameters constituting the model during fifth epidemic wave in Hong Kong (between 25th December, 2021 and 26th March 2022).** (A) Mobility index ($M_t$) (B) Daily average temperature ($T_t$).

**4 Partial data of BA.4/5 in Hong Kong (As of 15the September, 2022)**

| Date | Total cases | Predicted proportion for BA.4/5 | Cases for BA.4/5 | Predicted proportion for BA.2 | Cases for BA.2 |
|---|---|---|---|---|---|
| 2022/8/1 | 4019 | 4.40% | 177 | 90.40% | 3633 |
| 2022/8/2 | 3889 | 4.80% | 187 | 89.50% | 3481 |
| 2022/8/3 | 4321 | 5.30% | 229 | 88.60% | 3828 |
| 2022/8/4 | 4773 | 5.80% | 277 | 88.70% | 4234 |
| 2022/8/5 | 4223 | 6.60% | 279 | 86.70% | 3661 |
| 2022/8/6 | 4322 | 7.30% | 316 | 85.60% | 3700 |
| 2022/8/7 | 4035 | 8.20% | 331 | 84.70% | 3418 |
| 2022/8/8 | 3807 | 9.20% | 350 | 83.70% | 3186 |
| 2022/8/9 | 3783 | 10.00% | 378 | 83.00% | 3140 |
| 2022/8/10 | 4344 | 10.70% | 465 | 82.30% | 3575 |
| 2022/8/11 | 4180 | 11.60% | 485 | 81.50% | 3407 |
| 2022/8/12 | 4222 | 12.50% | 528 | 80.50% | 3399 |
| 2022/8/13 | 5148 | 14% | 721 | 79% | 4067 |
| 2022/8/14 | 4764 | 16.10% | 767 | 76.90% | 3664 |
| 2022/8/15 | 4699 | 17.80% | 836 | 74.80% | 3515 |
| 2022/8/16 | 4890 | 19.40% | 949 | 72.60% | 3550 |
| 2022/8/17 | 5563 | 20.90% | 1163 | 71.10% | 3955 |
| 2022/8/18 | 5811 | 22.40% | 1302 | 69.60% | 4044 |
| 2022/8/19 | 6260 | 24.50% | 1534 | 62.10% | 3887 |
| 2022/8/20 | 6210 | 26.20% | 1627 | 61.90% | 3844 |
| 2022/8/21 | 6276 | 28.00% | 1757 | 63.60% | 3992 |
| 2022/8/22 | 6380 | 29.60% | 1888 | 62.00% | 3956 |
| 2022/8/23 | 6410 | 31.40% | 2013 | 60.10% | 3852 |

| | | | | | |
|---|---|---|---|---|---|
| 2022/8/24 | 7677 | 33.60% | 2579 | 57.80% | 4437 |
| 2022/8/25 | 8282 | 35.90% | 2973 | 55.90% | 4630 |
| 2022/8/26 | 7665 | 38.30% | 2936 | 53.40% | 4093 |
| 2022/8/27 | 8225 | 41.10% | 3380 | 50.60% | 4162 |
| 2022/8/28 | 9495 | 43.80% | 4159 | 48.20% | 4577 |
| 2022/8/29 | 8252 | 46.50% | 3837 | 45.80% | 3779 |
| 2022/8/30 | 8611 | 49.10% | 4228 | 43.60% | 3754 |
| 2022/8/31 | 9267 | 51.50% | 4773 | 41.50% | 3846 |
| 2022/9/1 | 10342 | 53% | 5481 | 40.10% | 4147 |
| 2022/9/2 | 9714 | 54.70% | 5314 | 38.60% | 3750 |
| 2022/9/3 | 10222 | 56.30% | 5755 | 37.30% | 3813 |
| 2022/9/4 | 10490 | 58.30% | 6116 | 35.40% | 3713 |
| 2022/9/5 | 9869 | 60.70% | 5990 | 33.30% | 3286 |
| 2022/9/6 | 9187 | 62.50% | 5742 | 31.40% | 2885 |
| 2022/9/7 | 10030 | 64.10% | 6429 | 30% | 3009 |
| 2022/9/8 | 10910 | 65.40% | 7135 | 28.70% | 3131 |
| 2022/9/9 | 9922 | 67.10% | 6658 | 27.20% | 2699 |
| 2022/9/10 | 9602 | 68.70% | 6597 | 25.60% | 2458 |
| 2022/9/11 | 8904 | 70.30% | 6260 | 24.20% | 2155 |
| 2022/9/12 | 7772 | 71.60% | 5565 | 23.10% | 1795 |
| 2022/9/13 | 7067 | 73.80% | 5215 | 21.40% | 1512 |
| 2022/9/14 | 7148 | 75.20% | 5375 | 20.00% | 1430 |
| 2022/9/15 | 8023 | 76.60% | 6146 | 19.00% | 1524 |

**Table3 Daily reported case of BA.4/5 in Hong Kong[9]**

### Reference


1. https://www.covidvaccine.gov.hk/zh-HK/dashboard

2. https://www.med.hku.hk/en/news/press//-/media/HKU-Med-Fac/News/slides/20220314-sims_wave_5_omicron_2022_03_14_final.ashx

3. https://www.censtatd.gov.hk/tc/scode150.html

4. https://www.health.com/condition/infectious-diseases/coronavirus/what-is-incubation-period-omicron-covid-19

5. Thompson MG, Burgess JL, Naleway AL, et al. Prevention and Attenuation of Covid-19 with the BNT162b2 and mRNA-1273 Vaccines. N Engl J Med. 2021;385(4):320-329. doi:10.1056/NEJMoa2107058

6. Liang JB, Yuan HY, Li KK, et al. Path to normality: Assessing the level of social-distancing measures relaxation against antibody-resistant SARS-CoV-2 variants in a partially-vaccinated population. Comput Struct Biotechnol J. 2022;20:4052-4059. doi:10.1016/j.csbj.2022.07.048

7. https://ourworldindata.org/covid-google-mobility-trends

8. https://www.hko.gov.hk/tc/cis/climat.htm

9. https://www.dh.gov.hk/tc_chi/index.html
